# Polygenic Risk Score for Gastric Cancer

**DOI:** 10.1101/2020.08.11.20172221

**Authors:** Sharvani Padmaraj Pratinidhi, Saed Sayad

## Abstract

**Background:** Gastric Cancer is one of the most predominant types of cancer in the world, and its genomic links are currently being studied at great depth. In this paper, we work towards using Genome Wide Association Studies (GWAS) data for identifying the Single Nucleotide Polymorphisms (SNPs) which have the strongest correlation with the occurrence of gastric cancer through statistical tests and to leverage them to build a predictive model using machine learning algorithms. Polygenic risk scoring (PRS) is a straightforward predictive model for assigning genetic risk to individual outcomes (cancer or healthy).

**Method:** Genome Wide Association Studies (GWAS) data for Gastric Cancer was subjected to different statistical tests. Chi-square was used for feature selection by determining the degree of association between each probe (SNP) and the target (cancer or control). These results were used to eliminate many probes and proceed with only those that are statistically significant. Naïve Bayes Classifier and Catboost machine learning algorithms were used to build classification models to predict (score) gastric cancer.

**Results:** Naïve Bayes classifier and Catboost classification algorithms were used for modeling. The features were selected by performing Chi-square test on each of the 319283 SNPs in the data. These values were then ordered according to the negative log of the p-value and the top 5, 100 and 1000 features were used as inputs in the classification models. The Naïve Bayes classifier gave an accuracy in the range of 0.60 to 0.76 for different sets of features. The Catboost algorithm proved to be more suited for this application as it gave an accuracy above 0.90 for all subsets of features.

**Conclusions:** This paper aims at creating a highly accurate classification model to predict the occurrence of gastric cancer from GWAS genome data. The Catboost model with an input space of 100 SNPs yielded the best results with an accuracy of 0.93 and can be considered as a polygenic risk scoring model to score new patients for gastric cancer.

## Introduction

Gastric Cancer is one of the most commonly found type of cancer in the world and has a significantly high rate of mortality [1]. There is a good amount of research being done utilizing the data generated during surgery and chemotherapy to predict the survival of patients undergoing the treatment. Machine Learning models for classification like XGBoost and logistic regression have also been used to predict the occurrence of gastric cancer utilizing clinical test data and obtaining an accuracy of 0.89 [3].

Studies done in different parts of the world have shown that the genetic composition of a person plays an important role in determining their risk of having gastric cancer [1][5]. The National Institutes of Health (NIH) has repositories of genome-wide association study (GWAS) data which includes Single Nucleotide Polymorphism (SNP) level data of the entire genome [2]. This data is extremely useful to look for genetic markers that have a correlation with specific diseases.

The objective of this paper is to identify the SNPs having a strong correlation to the occurrence of gastric cancer by performing statistical tests and creating a machine learning model to predict it. This model can further be translated to obtain the Polygenic Risk score which expresses the relative risk of a disease in comparison with people of varied genetic composition [6]. We have used Naïve Bayes Classifier and Catboost machine learning algorithms to create a predictive classifier for gastric cancer and evaluated their performance on 5, 100 and 1000 SNPs as input features to obtain a model with high accuracy.

## Data

The Genomic Spatial Event (GSE) database on gastric cancer is based on an experiment for genome variation profiling by SNP array on a population from South Korea. It can be found on the Gene Expression Omnibus website: https://www.ncbi.nlm.nih.gov/geo/query/acc.cgiPacc-GSE58356

The data consisted of records of 1012 patients out of which 329 were diagnosed as gastric cancer cases and 683 were control cases. Each record had demographic details along with 319,283 SNPs. The SNPs were categorical variables with “AA”, “AB”, “BB” and “NC” as possible values for each. The target value was a binary variable where 1 stands for gastric cancer case and 0 stands for control case. The 319,283 SNPs were joined with the target vector and gender vector from the demographic data to form the complete dataset for this analysis.

**Figure 1:**
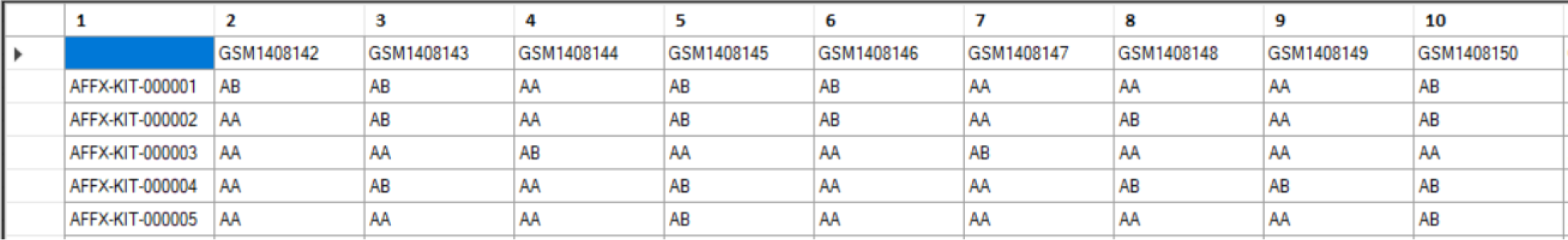
Snapshot of the dataset.

The data was further split into train (80%) and test (20%) sets for training and evaluation of the models. The data used for this analysis can also be found at: http://biomarkers.cs.rutgers.edu/prs/RNA/ugi.htm

## Exploration

Recent research has suggested that gastric cancer has strong genomic links. The GSE data contains records of control and gastric cancer cases expressed as 319,283 SNPs. The following statistical analysis was done to identify the top 5, 100 and 1000 SNPs with the strongest correlation to the occurrence of gastric cancer. These SNPs were further used to create a machine learning classification model to predict gastric cancer with high accuracy.

Figure 2 shows that the target variable contains 32% of gastric cancer cases and 68% of control cases. The gender variable is almost equally balanced with 51% males and 49% females.

**Figure 2:**
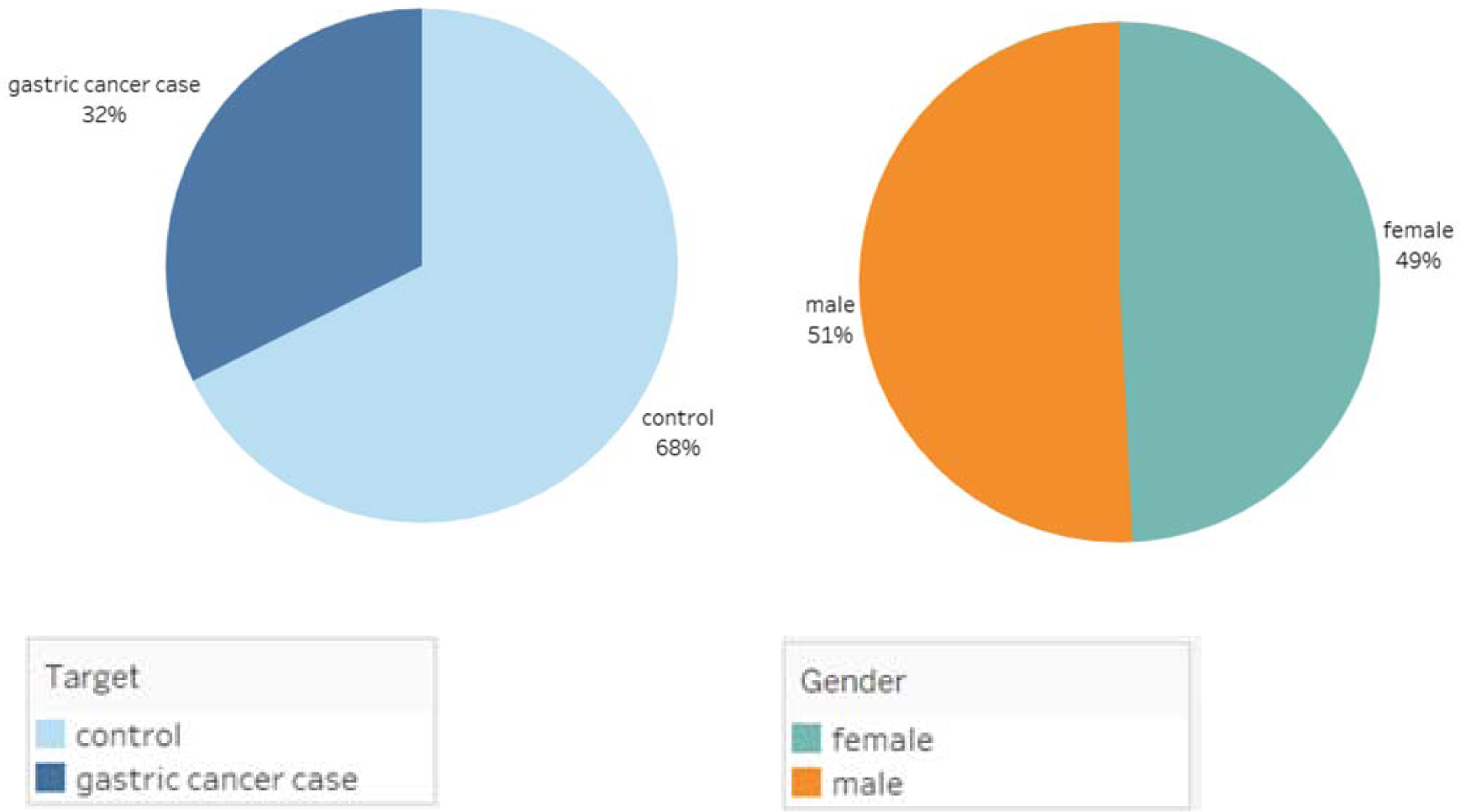
Univariate analysis of Target and gender variable.

The Chi-square test [4] was performed for each of the SNPs and the resulting p-value was used to calculate the significance of each SNP in predicting the target variable. The SNPs with the maximum significance were included in the models.

The negative log of the p-value is the significance level and it can be plotted against the chromosomes to visualize the significance of a chromosome in predicting gastric cancer. This plot is called the Manhattan Plot (Figure 3).

**Figure 3:**
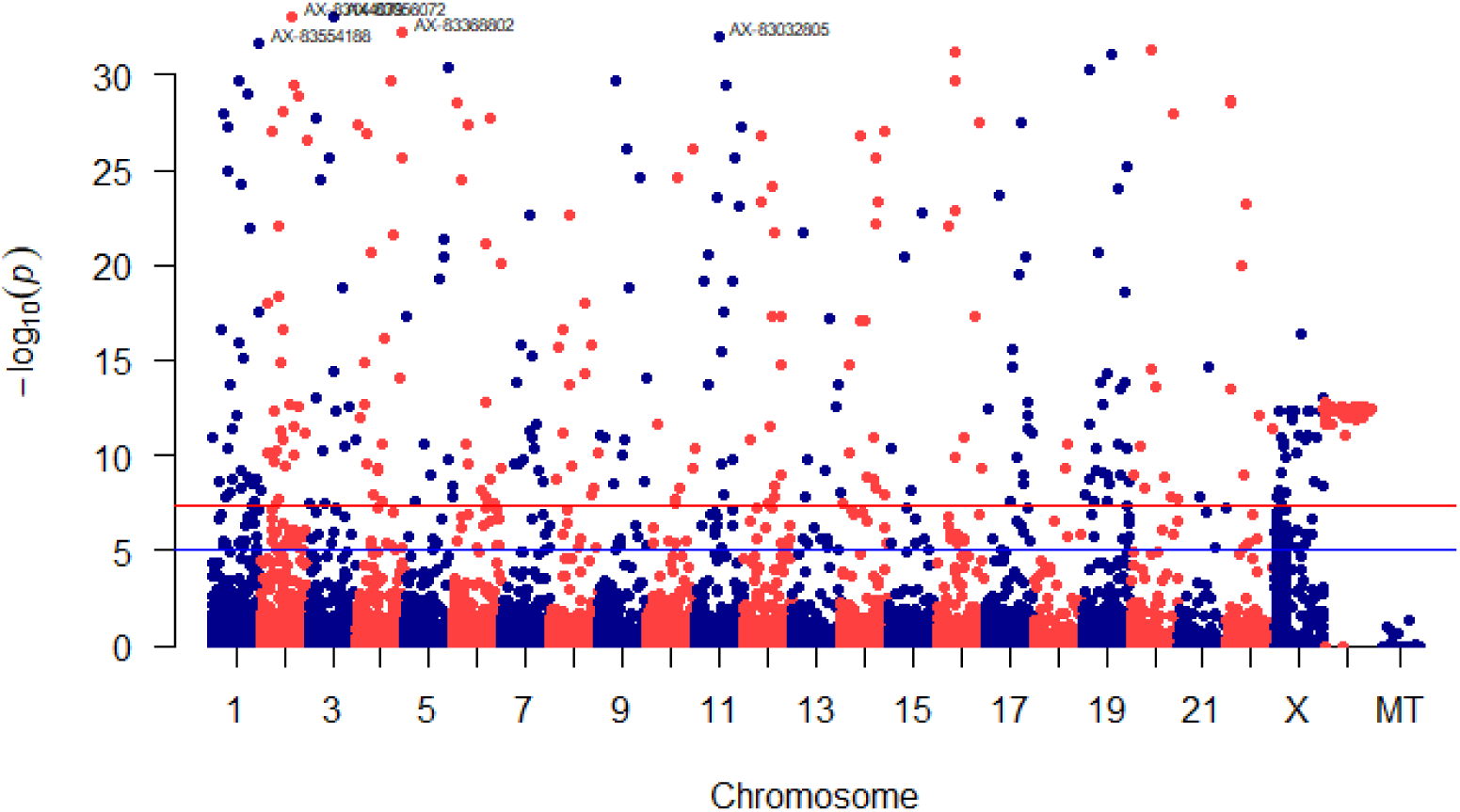
Manhattan Plot.

The top 5 SNPs were found (using Chi-square test) to be AX-83144079 in Chromosome 2, AX-83056072 in Chromosome 3, AX-83368802 in Chromosome 4, AX-83032805 in Chromosome 11 and AX-83554188 in Chromosome 1. Figure 4 shows an example of the Chi-square test done for SNP AX-83144079.

**Figure 4:**
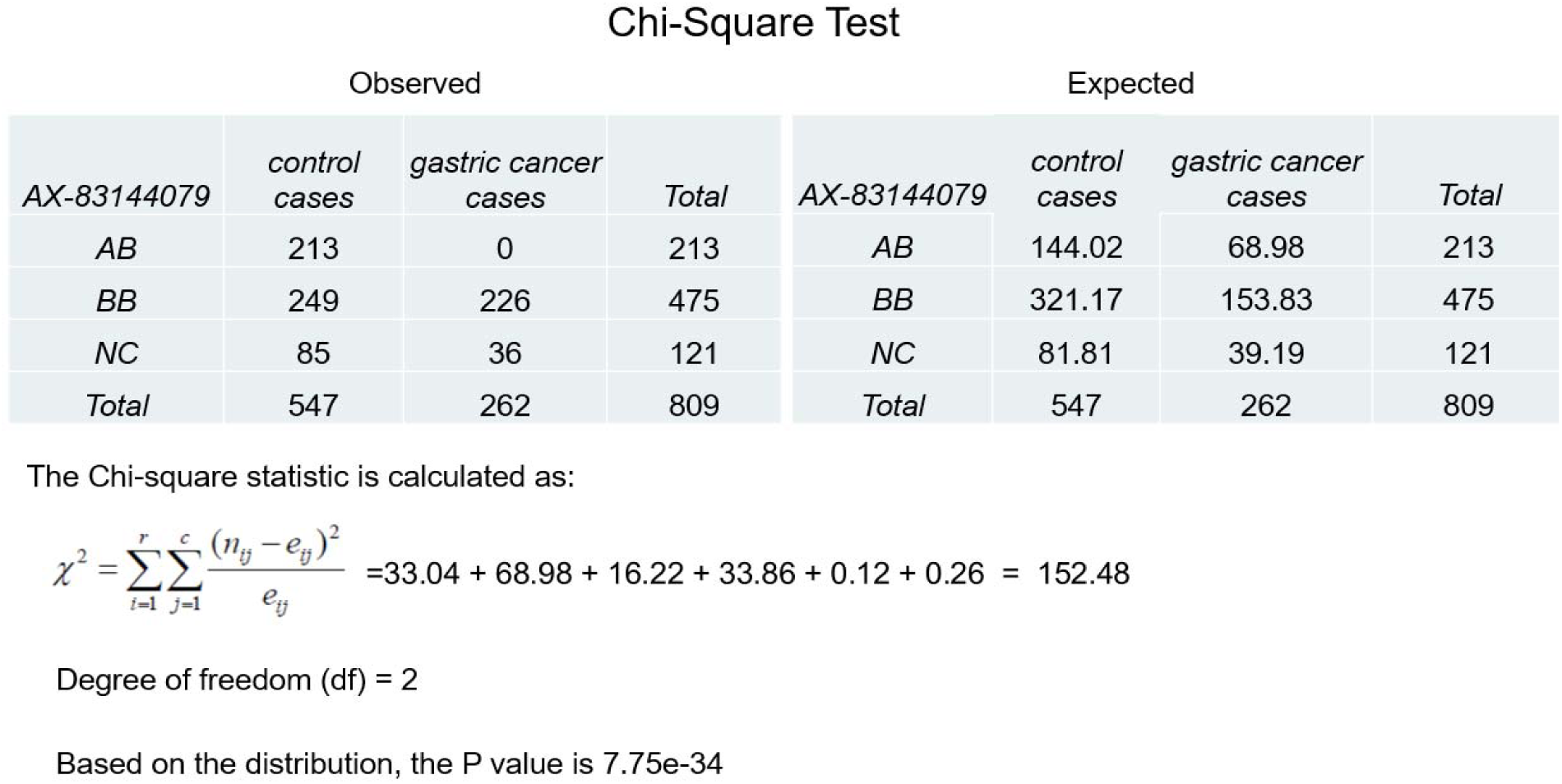
Chi-square Test.

### Modeling

Based on the Chi-square test, the top 5, 100 and 1000 out of the 319,283 SNPs were selected as input features to the Naïve Bayes and Catboost machine learning models and their performances were compared to each other. The data was split into training set and testing set in an 80:20 split.

The Naïve Bayes Classification model works on the assumption that all of the features are conditionally independent and contribute equally to the outcome [4]. The formula is given as:

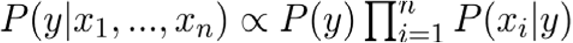

Catboost is a machine learning algorithm that uses gradient boosting on decision trees. This algorithm is well suited to handle the categorical nature of the data and thus yields good results.

The Catboost model with top 100 SNPs based was found to be the optimal model, as it gave a good level of accuracy while limiting the features used.

**Table 1:** Result of Catboost Model with 100 SNPs.

|  | Training | Testing |
| --- | --- | --- |
| Sensitivity | 0.96 | 0.91 |
| Specificity | 0.97 | 0.95 |
| Positive Predictive Value | 0.95 | 0.90 |
| Negative Predictive Value | 0.98 | 0.95 |
| **Accuracy** | **0.97** | **0.93** |

The results of for both Naïve Bayes and Catboost models are shown in the table below:

**Table 2:** Area Under Curve.

| Model | Train AUC | Test AUC |
| --- | --- | --- |
| Naïve Bayes with 5 SNPs | 0.74 | 0.72 |
| Naïve Bayes with 100 SNPs | 0.70 | 0.70 |
| Naïve Bayes with 1000 SNPs | 0.77 | 0.74 |
| Catboost with 5 SNPs | 0.73 | 0.71 |
| Catboost with 100 SNPs | 0.97 | 0.93 |
| Catboost with 1000 SNPs | 0.98 | 0.95 |

**Figure 5:**
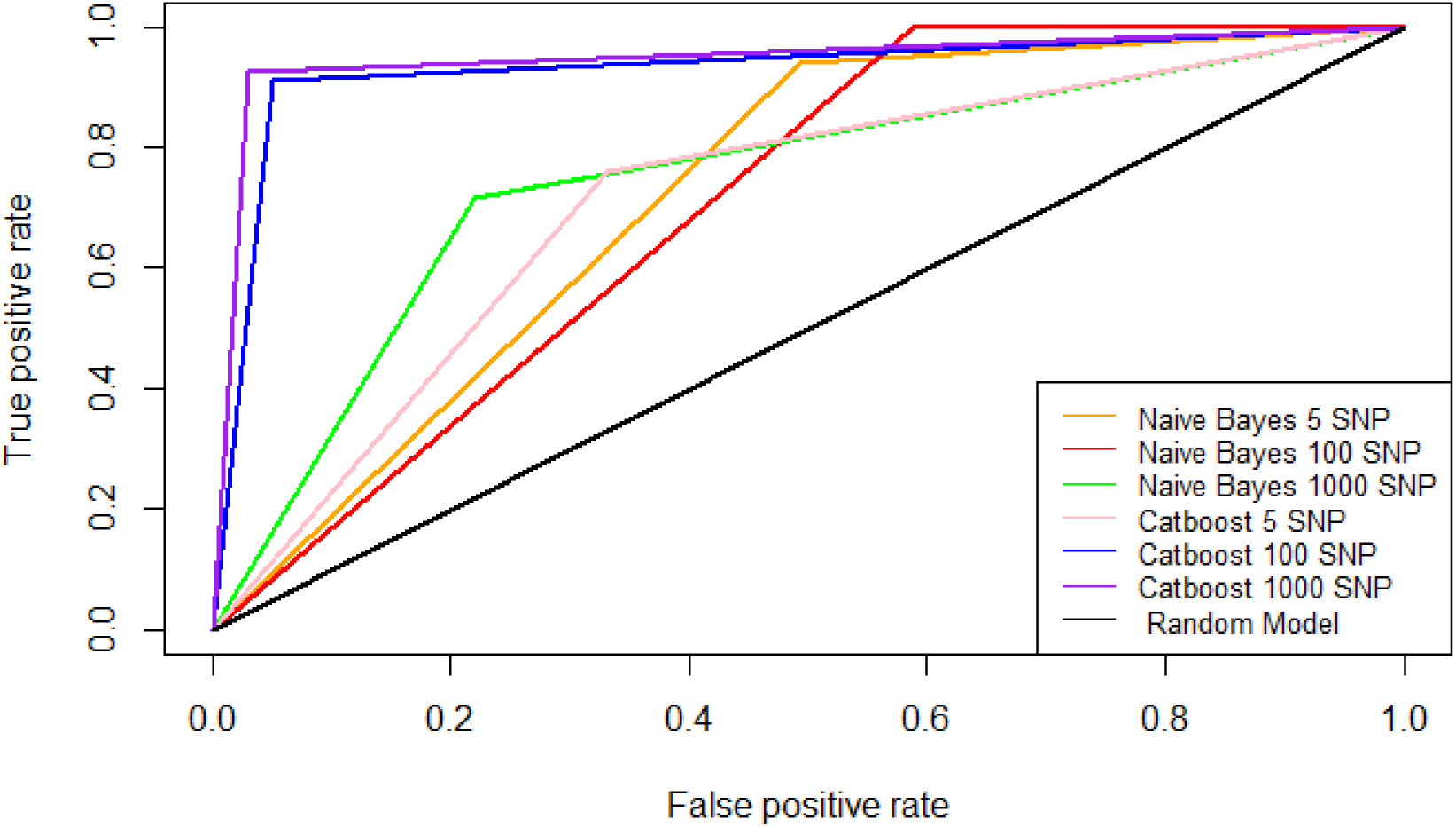
ROC Curve.

The ROC curve in Figure 5 shows the graph of sensitivity (True Positive Rate) vs specificity (False Positive Rate) [4] for all the models. We can observe that the Catboost model consistently yields better results as compared to the Naïve Bayes model and this corroborates with the overall accuracy of the models as

## Conclusions

In this paper we have attempted to identify the SNPs which have the strongest correlation with gastric cancer through statistical tests and used them to create a polygenic risk scoring model using machine learning algorithms. We have used two models, the Naïve Bayes classifier and Catboost classification algorithm and trained them on subsets of SNPs (5, 100 and 1000) to compare their performances. The top 5 SNPs were found to be AX-83144079, AX-83056072, AX-83368802, AX-83032805 and AX-83554188 in Chromosome 2, 3, 4, 11 and 1, respectively. The Naïve Bayes classifier gives moderate results as compared to the Catboost model which performs well even with just 5 input features. The optimal model is the Catboost model with an input space of 100 SNPs with an overall accuracy of 0.93 on the testing dataset. The selected model can be considered as a highly accurate Polygenic Risk Scoring model to score new patients for gastric cancer.

## Data Availability

The data is from the public domain and is available on the Gene Expression Omnibus Website https://www.ncbi.nlm.nih.gov/geo/query/acc.cgi?acc=GSE58356

https://www.ncbi.nlm.nih.gov/geo/query/acc.cgi?acc=GSE58356

## Notes

### Competing Interest Statement

The authors have declared no competing interest.

### Funding Statement

No external funding was received. This research was done as a capstone project.

### Author Declarations

This data is from the public domain and hosted on the Gene Expression Omnibus Website.

## References

1. Genome-Wide Association of Genetic Variation in the PSCA Gene With Gastric Cancer Susceptibility in a Korean Population https://pubmed.ncbi.nlm.nih.gov/30189721/

2. Dataset: https://www.ncbi.nlm.nih.gov/geo/auerv/acc.cgi?acc=GSE58356

3. Prediction of future gastric cancer risk using a machine learning algorithm and comprehensive medical check-up data: A case-control study https://www.nature.com/articles/s41598-019-48769-v

4. An Introduction to Data Science, https://www.saedsavad.com/data_mining_map.htm

5. Systematic evaluation of cancer-specific genetic risk score for 11 types of cancer in The Cancer Genome Atlas and Electronic Medical Records and Genomics cohorts https://www.ncbi.nlm.nih.gov/pmc/articles/PMC6558466/

6. Analysis of polygenic risk score usage and performance in diverse human populations https://www.nature.com/articles/s41467-019-11112-0

